# A circulating proteome-informed prognostic model of COVID-19 disease activity that relies on routinely available clinical laboratories

**DOI:** 10.1101/2022.11.02.22281834

**Authors:** William Ma, Antoine Soulé, Karine Tremblay, Simon Rousseau, Amin Emad

## Abstract

A minority of people infected with SARS-CoV-2 will develop severe COVID-19 disease. To help physicians predict who is more likely to require admission to ICU, we conducted an unsupervised stratification of the circulating proteome that identified six endophenotypes (EPs) among 731 SARS-CoV-2 PCR-positive hospitalized participants in the Biobanque Québécoise de la COVID-19, with varying degrees of disease severity and times to intensive care unit (ICU) admission. One endophenotype, EP6, was associated with a greater proportion of ICU admission, ventilation support, acute respiratory distress syndrome (ARDS) and death. Clinical features of EP6 included increased levels of C-reactive protein, D-dimers, interleukin-6, ferritin, soluble fms-like tyrosine kinase-1, elevated neutrophils, and depleted lymphocytes, whereas another endophenotype (EP5) was associated with cardiovascular complications, congruent with elevated blood biomarkers of cardiovascular disease like N-terminal pro B-type natriuretic peptide (NT-proBNP), Growth Differentiation Factor-15 (GDF-15), and Troponin T. Importantly, a prognostic model solely based on clinical laboratory measurements was developed and validated on 903 patients that generalizes the EPs to new patients recruited across all pandemic waves (2020-2022) and create new opportunities for automated identification of high-risk groups in the clinic. Thus, this novel way to address pathogenesis that leverages detailed phenotypic information but relies on routinely available information in the clinic to favor translation may find applications in other diseases beyond COVID-19.

## Introduction

An important challenge facing respirologists and critical care physicians is the heterogeneity in outcome following SARS-CoV-2 infections. A minority of people infected with SARS-CoV-2 will develop a severe form of coronavirus disease 2019 (COVID-19) requiring hospitalization and respiratory support. Defining the molecular mechanisms related to specific severe outcomes is important to identify treatable traits and improve the survival of critically ill patients. Successfully reaching this precision medicine goal requires a more granular definition of the underlying pathophysiology. A symptom-based method to discover molecular mechanisms of the disease is inherently confounded by the fact that the same higher-level condition, such as severe COVID-19 disease, can be produced by several different molecular mechanisms, a phenomenon termed the “many-one” limitation (1). Recent advances in computing strategies, such as machine learning, have enabled the development of methods that help overcome this limitation by, instead of using symptoms, starting from molecular profiles to define endophenotypes, i.e., subgroups of individuals who are inapparent to traditional classification methods but share a common set of molecular factors that can lead to identification of treatable traits (2-4). Current investigations of endophenotypes in COVID-19 have mainly relied on supervised approaches using fixed outcomes (such as disease severity) and integrating clinical variables at the onset (5). We hypothesize that using an unsupervised approach and exploiting a rich molecular dataset can provide novel mechanistic insights into the pathobiology of severe COVID-19 that can help physicians improve diagnosis, prognosis, and clinical management.

This study identified six endophenotypes linked to diverse clinical trajectories of COVID-19 using the extensive molecular phenotyping of a cohort of 731 SARS-CoV-2 positive hospitalized patients from the *Biobanque Québécoise de la COVID-19* (BQC19, www.quebeccovidbiobank.ca) (6), a prospective observational cohort of SARS-CoV-2-positive and negative participants recruited in the province of Québec, Canada, to improve our understanding of COVID-19 pathobiology and our capacity to alter disease outcomes. The molecular signature of each endophenotype was used to build a prognostic model of disease severity that generalizes the EPs to new patients and was validated on a separate group of 903 patients. This prognostic model solely utilizes clinical laboratory measurements, creating the possibility of automated identification of high-risk groups in the clinic.

## Results

### Unsupervised clustering of SARS-CoV-2-positive hospitalized BQC19 participants reveal endophenotypes associated with varying disease severity

We aimed to identify endophenotypes of COVID-19, based on the circulating proteome of patients, in a cohort of SARS-CoV-2 positive hospitalized participants in the BQC19 (Table 1) using an unsupervised approach. Figure S1 shows the distribution of the patient hospital admission dates and the corresponding waves of COVID-19 infection as defined by National Institute of Public Health of Quebec (INSPQ, https://www.inspq.qc.ca/covid-19). Consensus agglomerative clustering was performed on participants (n = 731, Table 1) for whom the circulating proteome was measured using a multiplex SOMAmer affinity array (SomaLogic, ∼5,000 aptamers) (7). The optimal number of clusters (k = 6) was identified first using two criteria: Akaike’s Information Criteria (AIC) and Bayesian Information Criteria (BIC) (Figure 1A). Then, consensus agglomerative clustering (Euclidean distance and Ward linkage) (8, 9) using 1000 bootstrap subsamples of the participants was performed to obtain six robust clusters (Figure 1, Figures S2 and S3). The distribution of Rand-Index, showing the concordance between each one of the 1000 subsampled clusterings and the final consensus clustering, is provided in Figure S2B (mean Rand-Index = 0.823), reflecting a high degree of consistency and robustness.

**Table 1:**
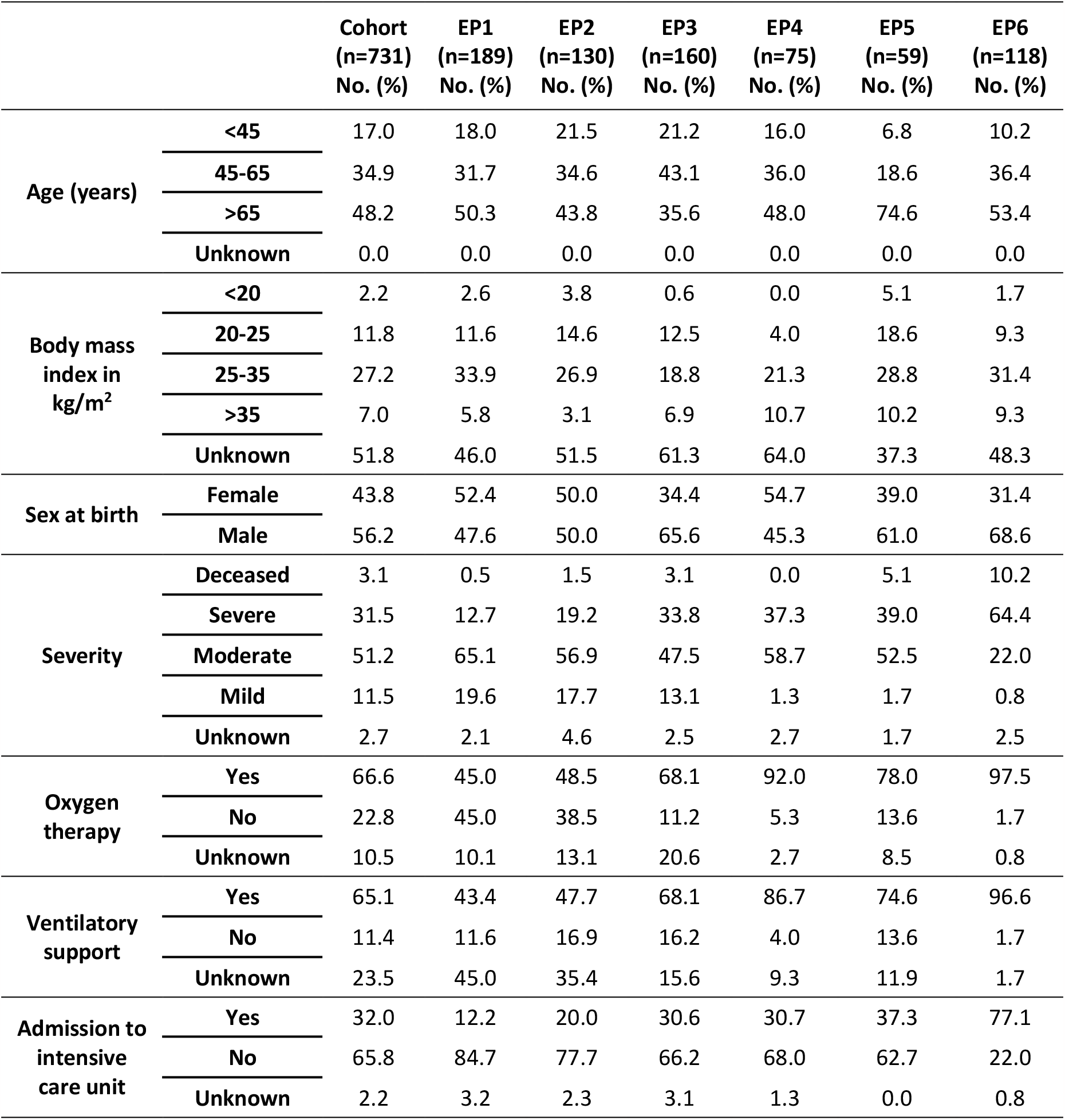
Clinical and pathological characteristics of the BQC19’s participants used in to identify endophenotypes (EPs) in this study.

**Figure 1:**
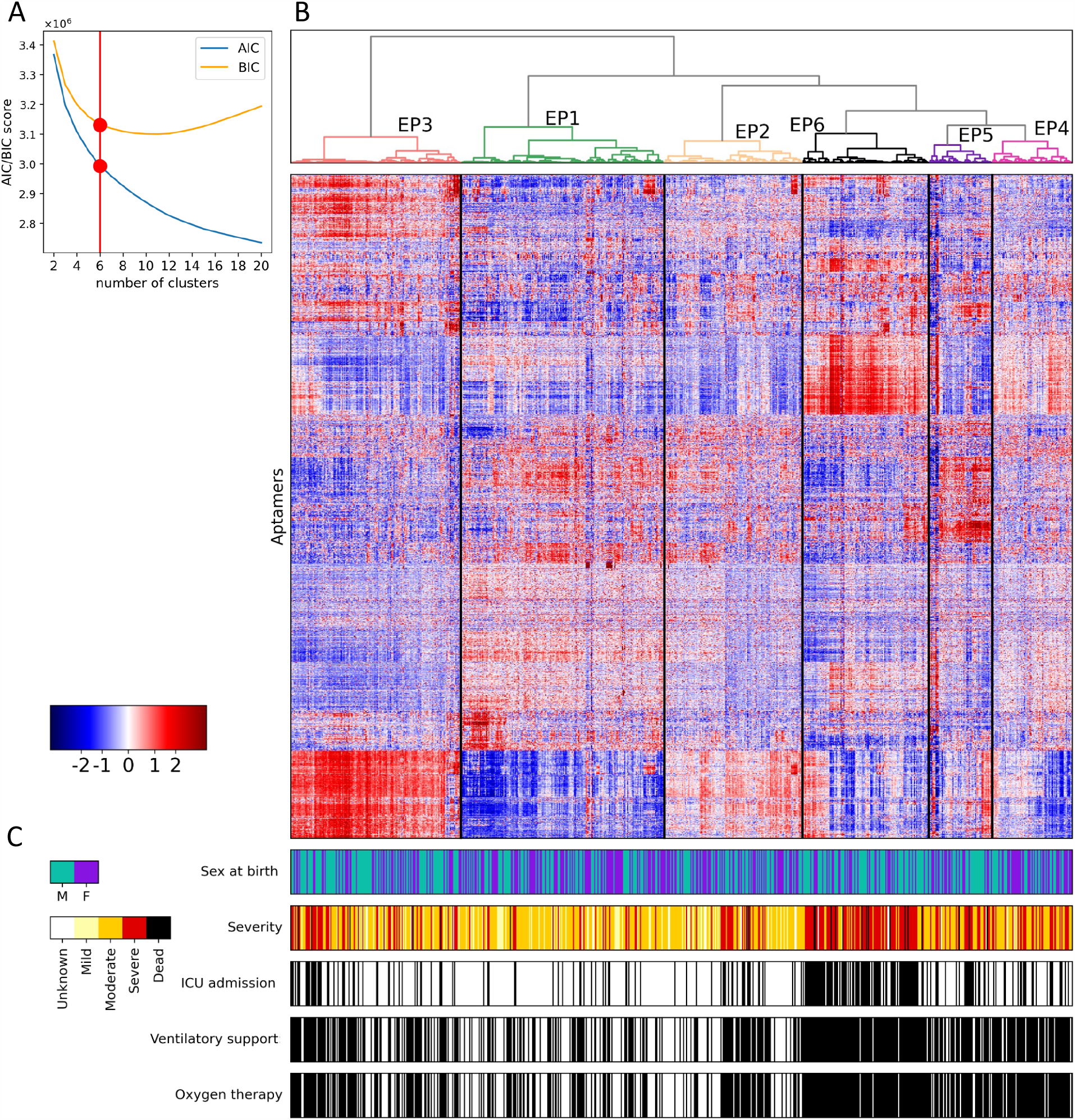
Unsupervised consensus clustering of SARS-CoV-2 positive patients. A) The elbow points (circles in red) of Akaike’s Information Criteria (AIC) and Bayesian Information Criteria (BIC) curves versus number of clusters consistently corresponded to k=6 as the optimal number of clusters. B) The heatmap shows the expression of aptamers (rows) in each sample (columns). The dendrogram shows the identified endophenotypes. C) Characterization of samples based on sex at birth, highest world health organization (WHO) severity level achieved, intensive care unit (ICU) admission, ventilatory support, and oxygen therapy. For the last three rows, a sample colored “black” reflects a label of “yes”.

The clinical and pathological characteristics of patients in each endophenotype is provided in Table 1. To characterize the identified endophenotypes (EPs) with respect to disease severity, we performed two-sided Fisher’s exact tests to assess their enrichment (or depletion) in either of “severe” or “deceased” outcomes. EP6 was significantly enriched in the severe/deceased outcomes (Benjamini–Hochberg false discovery rate (FDR) = 1.74E-21) with either of these outcomes observed in 74.6% of EP6 patients. Meanwhile, EP1 was significantly depleted in severe/deceased outcomes (FDR = 1.89E-13) (Figure 2A, Table 1, Table S1) with either of these outcomes observed in only 13.2% of EP1 patients. In addition, EP6 was enriched in participants (a) receiving oxygen therapy (FDR = 4.23E-18), (b) receiving ventilatory support (FDR = 4.59E-18), and (c) being admitted to intensive care unit (ICU) (FDR = 9.51E-28) (Figure 2A, Table 1, Table S1). Kaplan–Meier analysis (10) also confirmed that the identified EPs have a distinct temporal pattern of admission to ICU (multivariate logrank test P = 5.00E-30), with EP1 and EP6 having the highest and lowest probability, respectively, of not being admitted to ICU or dying prior to that in a 40-day span since their admission to the hospital (Figure 2C). A similar pattern was also observed when patients that died before admission to ICU were excluded (Figure S4, multivariate logrank test P = 5.39E-30). A two-sided Mann–Whitney U (MWU) test showed that patients in EP5 were generally older than other EPs (FDR = 7.73E-5), while EP3 included younger patients (FDR = 1.53E-4). Notably, EP6 (which had the most severe patients) did not show enrichment in older patients or individuals with high body mass index (BMI) (two-sided MWU FDR > 0.05) (Figures 2D-F, Table 1, Table S1).

**Figure 2:**
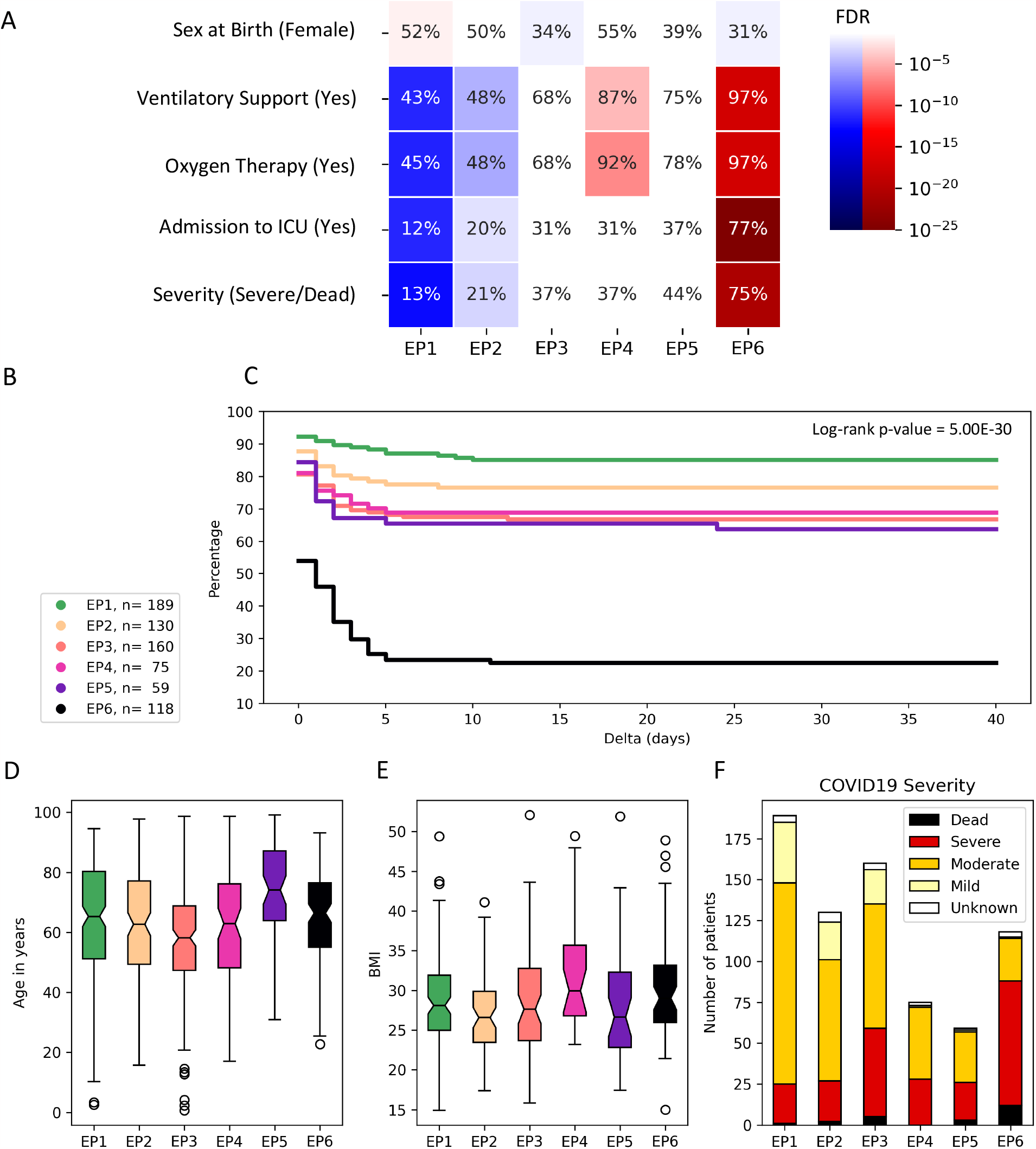
Characterization of endophenotypes (EPs). A) Enrichment or depletion of each EP in clinical variables (one cluster versus rest). Two-sided Fisher’s exact tests are used to calculate the p-values, which are corrected for multiple tests using Benjamini–Hochberg false discovery rate (FDR). Gradients of blue show depletion, while gradients of red show enrichment. FDR values above 0.05 are depicted as white. B) The number of patients in each EP and the colors used to represent them in panels C, D, and E. C) Kaplan– Meier analysis of the time between patients’ admission to the hospital and their admission to intensive care unit (ICU) (or death if earlier) for each EP (Delta). D) Distribution of age in each EP. E) Distribution of BMI in each EP. F) COVID-19 severity in each EP.

These analyses revealed that the unsupervised approach using the circulating proteome of the patients was able to identify endophenotypes with distinct disease characteristics and outcomes. We identified EP6 as a group of participants with an increase in key measures of COVID-19 disease severity, including admission to ICU and the need for ventilatory support.

### EP6 is enriched among BQC19 participants with acute respiratory distress syndrome

To better characterize all EPs with regards to different complications, we performed two-sided Fisher’s exact tests comparing each EP to the rest. In accordance with increased COVID-19 disease severity, EP6 was enriched in several medical complications including ARDS (FDR = 1.12E-11), acute kidney injury (FDR = 5.73E-8), secondary bacterial pneumonia (FDR = 2.25E-5), liver dysfunction (FDR = 1.37E-3), cardiovascular complications (FDR = 1.37E-3), and bacteremia (FDR = 4.28E-3) (for the full list, see Figure 3 and Table S2). Notably, the frequency of ARDS was 9% in EP1 compared to 50% in EP6 making this complication a key feature of this endophenotype (Figure 3, Table S2).

**Figure 3:**
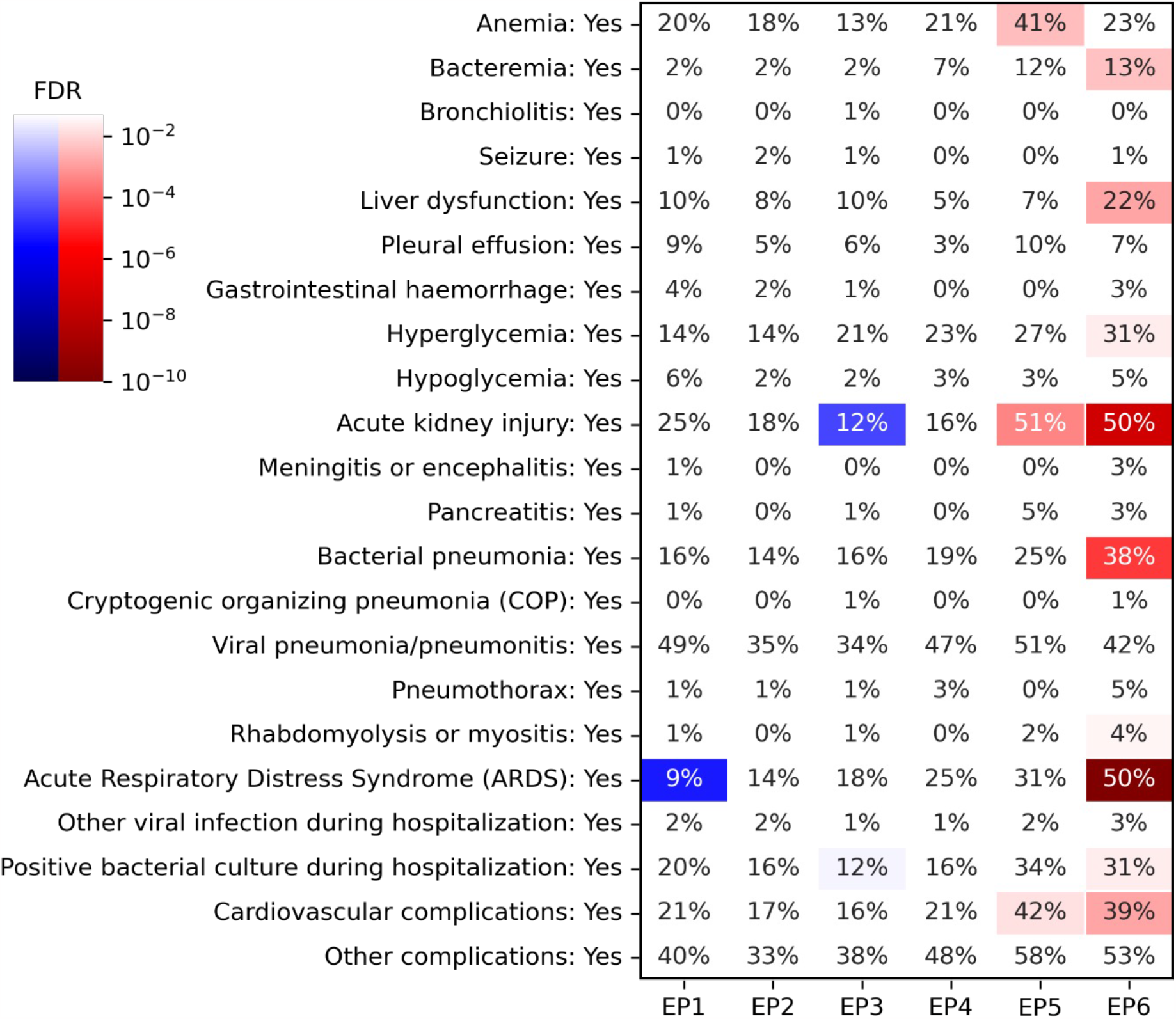
Frequency and significance of complications in different EPs. The value in each cell shows the percentage of patients of that EP (column) that suffered from the complication (row). The colors represent two-sided Fisher’s exact test false discovery rate (FDR, corrected for multiple tests). Red represents enrichment, while blue represents depletion. FDR values below 0.05 are shown as white.

### Clinical laboratories reveal that members of EP6 have increased levels of C-reactive protein, D-dimers, elevated neutrophils, and depleted lymphocytes

To further characterize each EP, we assessed the clinical laboratory results obtained from blood draws and compared them between the groups. We focused on 21 markers that were measured in at least 50% of the patients of the cohort and used the summary value reported in the BQC19 database corresponding to the most extreme measurement among multiple blood draws (Figure 4A, Table S3 also includes first blood draw characteristics). Figure 3A shows the elevation and depletion of these markers in the identified EPs. EP6 is characterized by abnormal values in markers of inflammation (lymphopenia, total white blood cell count, neutrophilia, C-reactive protein (CRP)), liver damage (alanine aminotransferase (ALT)), coagulopathy (D-dimers, low hemoglobin, International Normalized Ratio (INR), and hyperglycemia (glucose). We also used 22 markers from the Elecsys diagnostic panel (Roche Diagnostic) to further characterize EP6, (Figure 4B and Table S3). This led to additional elevated and highly significant markers: (a) alpha-1 antitrypsin, an acute phase reactant elevated during inflammatory conditions; (b) Interleukin 6 (IL-6), a pleiotropic cytokine associated with systemic inflammatory response syndrome (11), shown to be elevated in severe COVID-19 (12) and linked to endothelial damage and liver injury (13); (c) ferritin, an iron-storage protein and acute phase reactant, elevated in COVID-19, and like other hyperferritinic syndrome, associated with coagulopathy (14, 15); and (d) soluble vascular endothelial growth factor (VEGF) receptor sFLT1 (soluble fms-like tyrosine kinase-1), previously shown to be associated with endothelial damage and COVID-19 severity (16). The overall characteristics of each EP are summarized in Table 2.

**Table 2:** Summary of the characteristics of each endophenotype. In this table, High (Low), denoted as H (L) implies that the average value of the variable in the corresponding EP was significantly higher (lower) than the other EPs (considered together), while N (Nondescript) implies that it was not significantly different.

| Endophenotype | Age | Sex at birth | BMI | Blood markers |
| --- | --- | --- | --- | --- |
| EP1 | H | F | N | High lymphocyte,<br>Low neutrophil |
| EP2 | N | N | L | High albumin,<br>Low white blood cells |
| EP3 | L | M | N | High platelet,<br>Low creatinine |
| EP4 | N | N | N | High lactate |
| EP5 | H | N | N | High creatinine,<br>Low haemoglobin |
| EP6 | N | M | N | High white blood cells,<br>Low lymphocyte |

Abbreviations used: H = High, L = Low, N = Nondescript, F = Female, M = Male

**Figure 4:**
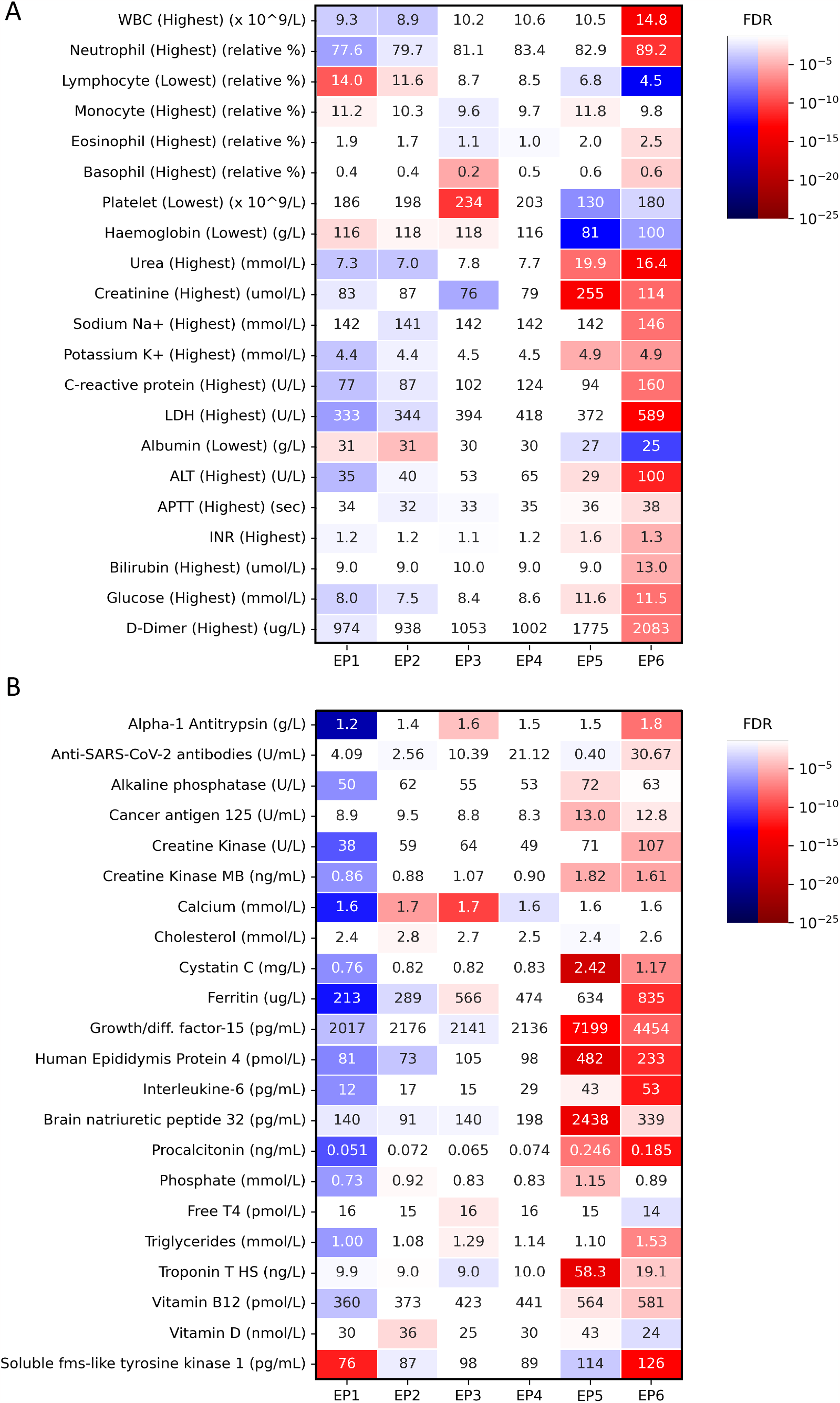
Patterns of blood markers and Roche diagnostic markers in the endophenotypes (EPs). Heatmaps show the false discovery rate (FDR) values for two-sided one-vs-rest Mann–Whitney U tests for 21 blood markers (most extreme value during hospitalization) (A) and 22 Roche diagnostic markers (B) for each EP. FDR values below 0.05 are shown as white. The numerical values show the median value of the maker in each EP. Abbreviations used: WBC = white blood cells, LDH = lactate dehydrogenase, ALT = alanine aminotransferase, aPTT = activated partial thromboplastin time, INR = International Normalized Ratio.

### EP5 is associated with cardiovascular complications

EP5 comes second in the order of severity established in Figure 2. Interestingly, it is molecularly and clinically distinct from EP6 (Figures 2-4, Table 1). A striking feature of EP5 is the increase in markers of cardiovascular diseases, such as higher levels of N-terminal pro B-type natriuretic peptide (NT-proBNP), indicative of ventricular dysfunction (17), Growth Differentiation Factor-15 (GDF-15) associated with cardiometabolic risk (18) and Troponin T linked to cardiac damage all suggestive of high risk for cardiovascular events (19) (Figure 4B). Accordingly, this group was enriched for cardiovascular complications during hospitalization (FDR = 1.46E-2, Figure 3). As postulated, the unsupervised clustering was able to distinguish different types of COVID-19 disease trajectory.

### A computational prognostic model based on blood biomarkers predicts EPs in a separate validation cohort

Since each EP showed a clear and distinct clinical laboratory result signature based on 21 blood markers and 22 Elecsys diagnostic markers, we sought to develop a computational prognostic model of disease severity based on these signatures. We focused on data from the first blood draw (Figure 4B and S5, Table S3) and developed a nearest-centroid classifier, capable of dealing with missing values, to predict EPs based on these 43 markers (see Methods for details). To test the prognostic ability of this model on an independent yet similar dataset, we analyzed 903 SARS-CoV-2 positive hospitalized BQC19 participants that did not have circulating proteome data and had not been used to identify the endophenotypes (see Figure S6 for the distribution of the patient hospital admission dates). These patients were recruited across all waves of the pandemic between March 2020 and October 2022. The clinical and pathological characteristics of patients in each predicted endophenotype (PEP) are provided in Figure 5 and Table S4.

**Figure 5:**
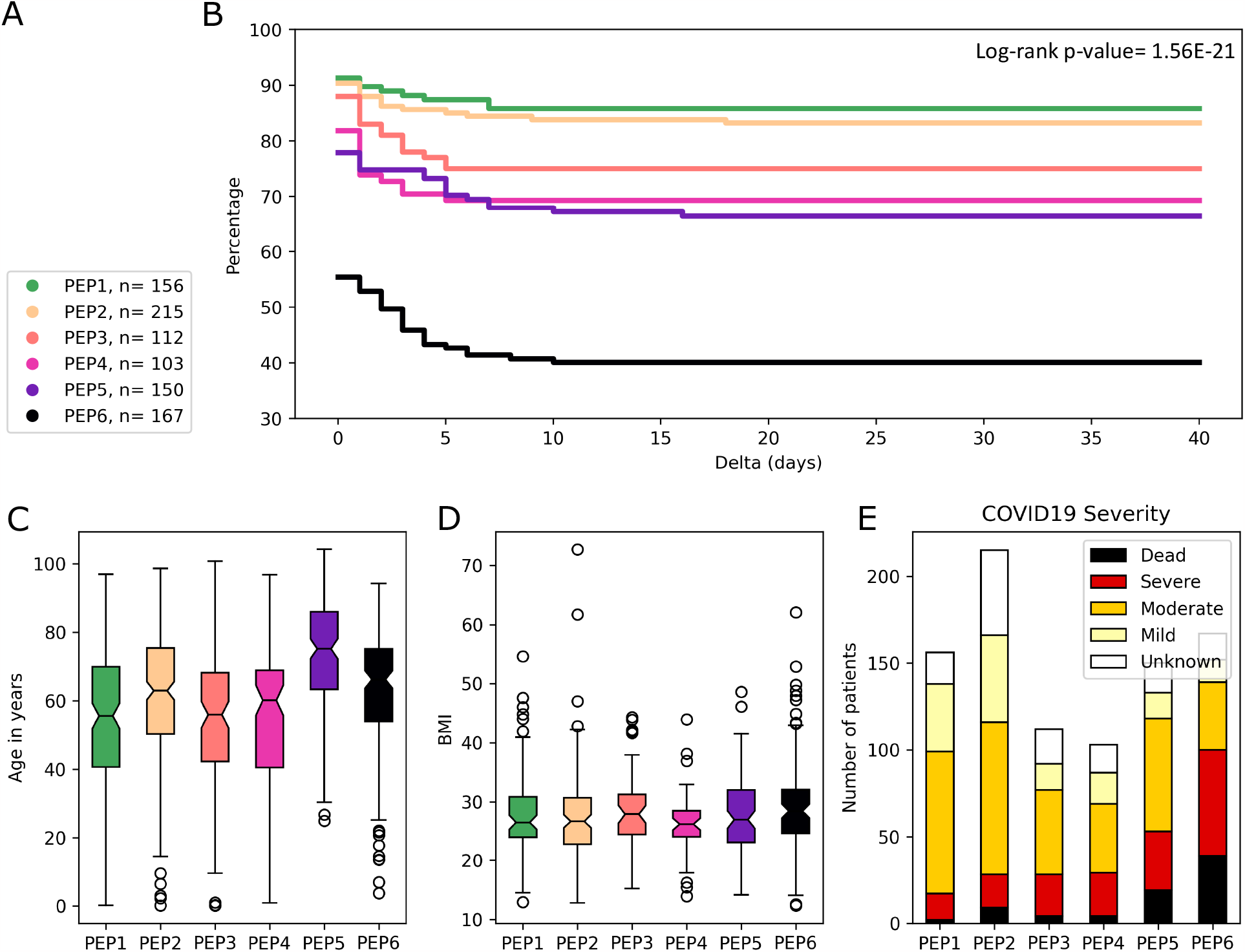
Characterization of predicted endophenotypes (PEPs) based on the prognostic model using 21 blood markers and 22 Roche diagnostic markers. A) The number of patients in each PEP and the colors used to represent them in panels B, C, D, and E. C) Kaplan–Meier analysis of the time between patients’ admission to the hospital and their admission to intensive care unit (ICU) (or death if earlier) for each PEP (Delta). D) Distribution of age in each PEP. E) Distribution of BMI in each PEP. F) World health organization COVID-19 severity in each PEP.

Our prognostic model identified 167 of these 903 patients as belonging to predicted EP6 (PEP6). Fisher’s exact tests showed significant enrichments of PEP6 in severe/deceased (FDR = 5.62E-21), while PEP1 and PEP2 were significantly depleted in these outcomes (FDR = 5.29E-8 and FDR = 1.19E-8, respectively), as shown in Table S4. Like EP6, PEP6 was also significantly enriched in participants (a) receiving oxygen therapy (FDR = 2.87E-13), (b) receiving ventilatory support (FDR = 1.50E-13), and (c) being admitted to ICU (FDR = 1.34E-19) (Table S4). Kaplan–Meier analysis also confirmed that these PEPs have a distinct temporal pattern of admission to ICU (multivariate logrank test P = 1.56E-21), with PEP6 having the highest chance of being admitted to ICU (or dying prior to that) in the 40-day span following admission to hospital (Figure 5B). These results suggest that our prognostic model based on 43 blood biomarkers can be used to generalize the definition of endophenotypes to patients for whom proteomic data is not available. Since the 21 blood markers are more commonly available, we also developed a prognostic model only based on these markers, which also showed strong prognostic capabilities (Figure S7). Therefore, it is possible to leverage detailed molecular information on a smaller number of participants to predict clinical outcomes on a larger population using routinely available information collected during hospitalization.

## Discussion

In this study, we have bridged the gap between the circulating proteome and routinely available blood diagnostic biomarkers, using machine learning algorithms, to prognosticate COVID-19 outcomes in hospitalized patients. The model performed on participants recruited across all the pandemic waves from 2020 to 2022, demonstrating that it performs despite mutations in infecting strains and the development of immunity. This showcases a novel analytical pipeline that can support physicians in making more informed decision on potential unfavorable trajectories early during hospitalization and adjust follow-ups and treatments accordingly.

The major strength of this study is the use of an unsupervised approach for analysis of a large and well-phenotyped cohort. This broad-based approach led to the identification of six COVID-19 disease endophenotypes in hospitalized patients that could not be captured by simply classifying the population solely based on severity, with different clinical trajectories and distinguishing characteristics that are summarized in Tables 1 and 2. We identified two endophenotypes with more favorable outcomes (EP1 and EP2), three endophenotypes with intermediate outcomes in terms of severity (EP3, EP4 and EP5) and one endophenotype which led to worst outcomes compared to all others (EP6). EP6, was associated with ARDS, the worst clinical manifestation of COVID-19 that was reflected by a greater proportion of ICU admission, mechanical ventilation, and severe/fatal outcomes (Figures 2 and 3). Clinical features of this endophenotype were consistent with published literature, including increased levels of CRP, D-dimers, IL-6, ferritin, sFLT1, elevated circulating neutrophils, and reduced peripheral blood lymphocytes (Figure 3, Table S3), presenting a profile associated with systemic inflammatory response syndrome and abnormal coagulation. Possible molecular effectors of COVID-19 disease severity in EP6 are discussed in an accompanying study. Another endophenotype (EP5), while leading to unfavorable clinical trajectory during hospitalization, was instead associated with clear markers of cardiovascular disease, cardiovascular complications during hospitalization, and older age. The distribution of clinical laboratories in each endophenotype was sufficient to train an accurate prognostic model that could readily support future clinical care, since it only requires data from routine clinical laboratory results for prognosis.

The identification of endophenotypes was done systematically using robust consensus clustering of aptamer expression levels in which the optimum number of clusters was determined congruently using two well-established measures: AIC and BIC. The consensus clustering using bootstrap sampling (1000 times) ensured identification of robust clusters that are not sensitive to exclusion of some of the samples (20% randomly selected and excluded at each cycle). The mean Rand-index between each of the 1000 subsampled clusterings and the final consensus clustering was 0.823, reflecting a high degree of concordance and robustness. Moreover, identifying the best number of clusters using AIC/BIC (both of which agreed with each other) allowed us to reveal the patterns of the EPs directly from the data instead of imposing a pattern onto it through human supervision. This is an important strength of the study that enabled us to identify distinct molecular patterns of patients that could have remained undetected using other traditional approaches.

Additionally, to improve the translational applicability of the EPs, we developed a prognostic model based only on measurements of conventional clinical laboratory blood markers to test the generalizability of these endophenotypes to samples without measured aptamer data. Characteristics of EPs predicted solely based on their blood markers were consistent with the original EPs, suggesting that clinical blood markers could be used as surrogates for assignment of these EPs to new patients and potentially automating identification of high-risk groups in the clinic. This approach takes into account the effect of multiple blood variables simultaneously and incorporates the full distribution of each variable. This is in contrast to the clinical laboratory results that are automatically flagged as within or outside normal range, one variable at a time, therefore increasing the clinical applicability of our model by leveraging a wider spectrum of information to prognosticate patient outcomes.

### Limitations and considerations

The data presented in this study comes from individuals participating in the BQC19, a prospective observational cohort built to study COVID-19 in Québec (Canada) with its specific population profile as reported previously (6). A chronological bias may also be present, as most of the participants used for endophenotyping in this study were recruited during the first two waves of the pandemic (Figure S1), prior to widespread vaccination in Québec and the appearance of the Omicron variant and sub-variants. Therefore, some of the features of the identified endophenotypes may change over the course of the pandemic. It will be essential to continue longitudinal assessments of the molecular profiles to better understand the dynamic nature of host-pathogen interactions. It will also be interesting to compare the profiles of COVID-19 ARDS to other viral-induced ARDS, to identify common as well as distinguishing features of these conditions.

## Conclusion

Respiratory infections represent an important challenge for respirologists and critical care physicians due to the heterogeneity of outcomes. Developing better ways to prognosticate poor outcomes is crucial in improving patients’ care and survival. In this manuscript, we proposed a novel experimental approach that leverages detailed proteomic information but relies on routinely available information in the clinic for prognostication to favor translation that may find applications in many other diseases beyond COVID-19.

## Methods

### Datasets and preprocessing

The Biobanque Québécoise de la COVID-19 (BQC19; www.quebeccovidbiobank.ca) is aimed at coordinating the collection of patients’ data and samples for COVID-19 related research. Data and samples were collected from ten sites across the province of Québec (Canada) (6). BQC19 organizes the collected data, including clinical information and multi-omics experimental data, before making it available in successive releases. For this study, we used the circulating proteome determined using SOMAmers. Our main corpus of analysis consisted of n = 1,634 hospitalized and SARS-CoV-2 positive patients (based on qRT-PCR) of BQC19. This included n = 731 patients (Figure S1) for which both clinical and proteomic data was available as well as n = 903 patients (Figure S6) for whom proteomic data was not available but whose clinical data contained measurements for more than half (at least 11 out of 21) of the blood markers that we used as a validation set for the prognostic model developed in this study.

We also obtained data (n = 731) corresponding to the circulating proteome measured between April 2, 2020 and April 20, 2021 by a multiplex SOMAmer affinity array (SomaLogic, 4,985 aptamers) from BQC19. When measurements of the same patients but at different time points were available, we used the one corresponding to the first time point. SomaScan is a biotechnological protocol commercialized by the SomaLogic company (7). It relies on a set of artificial aptamers linked to a fluorophore and each designed to bind a single protein. Once added to the sample, the activity of each aptamer is measured through fluorescence and used to approximate the expression level of the targeted protein. SomaScan protocol comprises several levels of calibration and normalization to correct technical biases. Log2 and Z-score normalization were performed on each aptamer separately in addition to the manufacturer’s provided normalized data (hybridization control normalization, intraplate median signal normalization, and median signal normalization). Since the data was analyzed by SomaLogic in two separate batches, we applied the z-score transformation separately to each batch, to reduce batch effects. These additional transformations ensure that the measured values of different aptamers are comparable and can be used in cluster analysis.

### Consensus agglomerative clustering

Patients were clustered using agglomerative clustering with Euclidean distance and Ward’s linkage (8, 9). To identify number of clusters k, we used the elbow method based on the AIC and BIC. More specifically, we calculated the AIC and BIC for clustering using k = 2, 3, …, 20 and used the Kneedle algorithm (20) to identify the value of k corresponding to the “elbow”, where increasing the value of k does not provide much better modeling of the data. Kneedle identified k = 6 as the number of clusters based on both AIC and BIC (Figure 1A).

Given the number of clusters in the data, we then used consensus clustering with sub-sampling to obtain robust endophenotypes. We randomly sampled 80% of the patients 1000 times. Each time, we used the agglomerative clustering above with k = 6 to identify clusters. Given these 1000 clusterings, we calculated the frequency of two patients appearing in the same cluster, when both were present in the randomly formed dataset. We then performed one final agglomerative clustering of these frequency scores to identify the six endophenotypes (Figure S2A and Figure 1B).

### Nearest-centroid predictor based on blood markers

In order to predict endophenotypes from blood tests, we developed a missing-value resilient nearest-centroid classifier. We used the dataset of patients that were used to form the original EPs (n = 731) as the training set and the dataset of patients that did not have proteome data as the validation set (n = 903). First, we z-score normalized each of the 43 markers across all the patients in the training set, one marker at a time. We then formed a marker signature (a vector of length 43) for each EP. Each element of an EP’s signature corresponds to the mean of the corresponding marker across all patients of that EP.

To predict the EP label of each patient in the test set, we first z-score normalized their blood marker measurements using the mean and standard deviation of the markers calculated from the training set. Then, we calculated the cosine distance between each test patient’s blood marker profile and the centroids (excluding missing values) and identified the nearest EP as the predicted EP (PEP) label of the patient.

### Statistics

Several non-parametric tests, including Mann–Whitney U test, Fisher’s exact test, and Spearman’s rank correlation, were used in this study. Benjamini–Hochberg false discovery rate (FDR) was used to adjust the p-values for multiple tests.

### Study approval

The study was approved by the Institutional Ethics Review Board of the “Centre intégré universitaire de santé et de services sociaux du Saguenay-Lac-Saint-Jean” (CIUSSS-SLSJ) affiliated to the Université de Sherbrooke [protocol #2021-369, 2021-014 CMDO – COVID19].

## Supporting information

STROBE Checklist

## Data Availability

Input data corresponding to the cohort can be obtained form BQC19 (www.quebeccovidbiobank.ca). The data generated as a result of the analyses are provided as tables and supplementary tables.

## Acknowledgements

This work was made possible through open sharing of data and samples from the Biobanque Québécoise de la COVID-19, funded by the Fonds de recherche du Québec - Santé, Génome Québec, the Public Health Agency of Canada and, as of March 2022, the Ministère de la Santé et des Services Sociaux du Québec. We thank all participants to BQC19 for their contribution. This study was supported by the Fonds de recherche du Québec - Santé (FRQS)-Cardiometabolic Health, Diabetes and Obesity Research Network (CMDO)-Initiative. This work was also supported by Natural Sciences and Engineering Research Council of Canada (NSERC) grant RGPIN-2019-04460 (AE). The authors also acknowledge the in-kind contribution of Roche Diagnostics, a division of Hoffmann–La Roche Limited, which provided the reagents for the biomarker analyses conducted on the BQC19 blood samples.

