## Supplementary File S1 for "A circulating proteome-informed prognostic model of COVID-19 disease activity that relies on routinely available clinical laboratories"

### Supplementary Figures

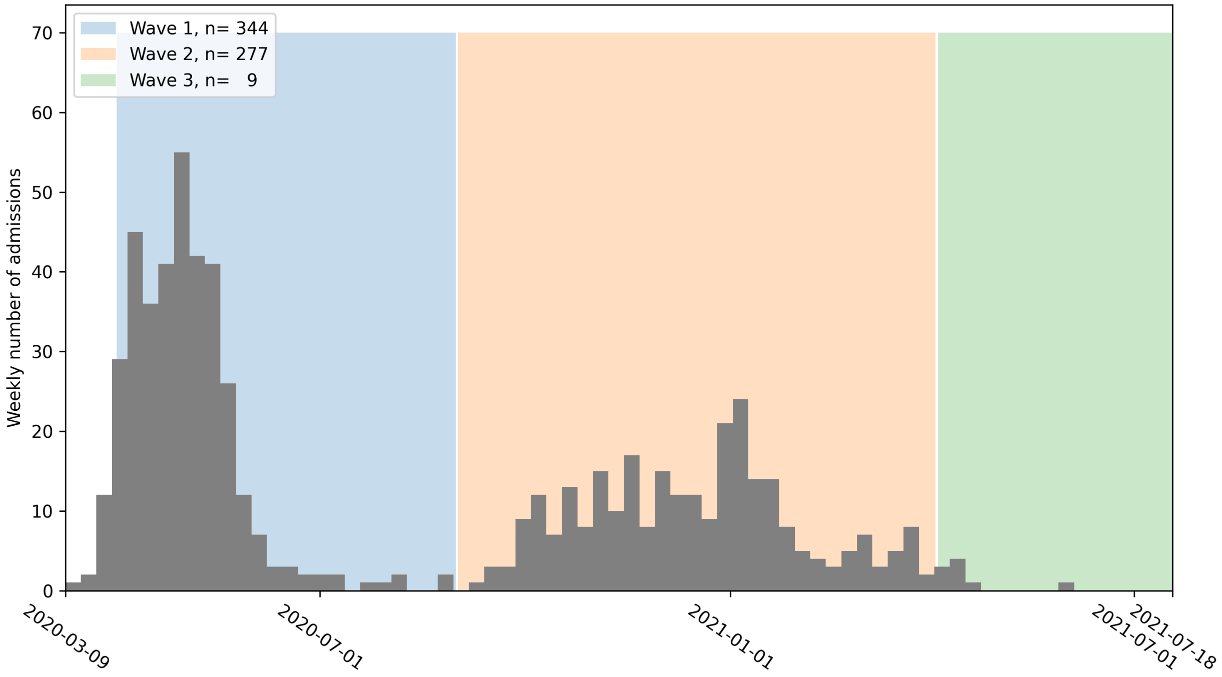


**Figure S1:** Weekly histogram of hospital admissions for patients used to define the endophenotypes (EPs) with known admission dates. Colors show different waves of COVID19 pandemic in Quebec according to National Institute of Public Health of Quebec (https://www.inspq.qc.ca/covid-19).

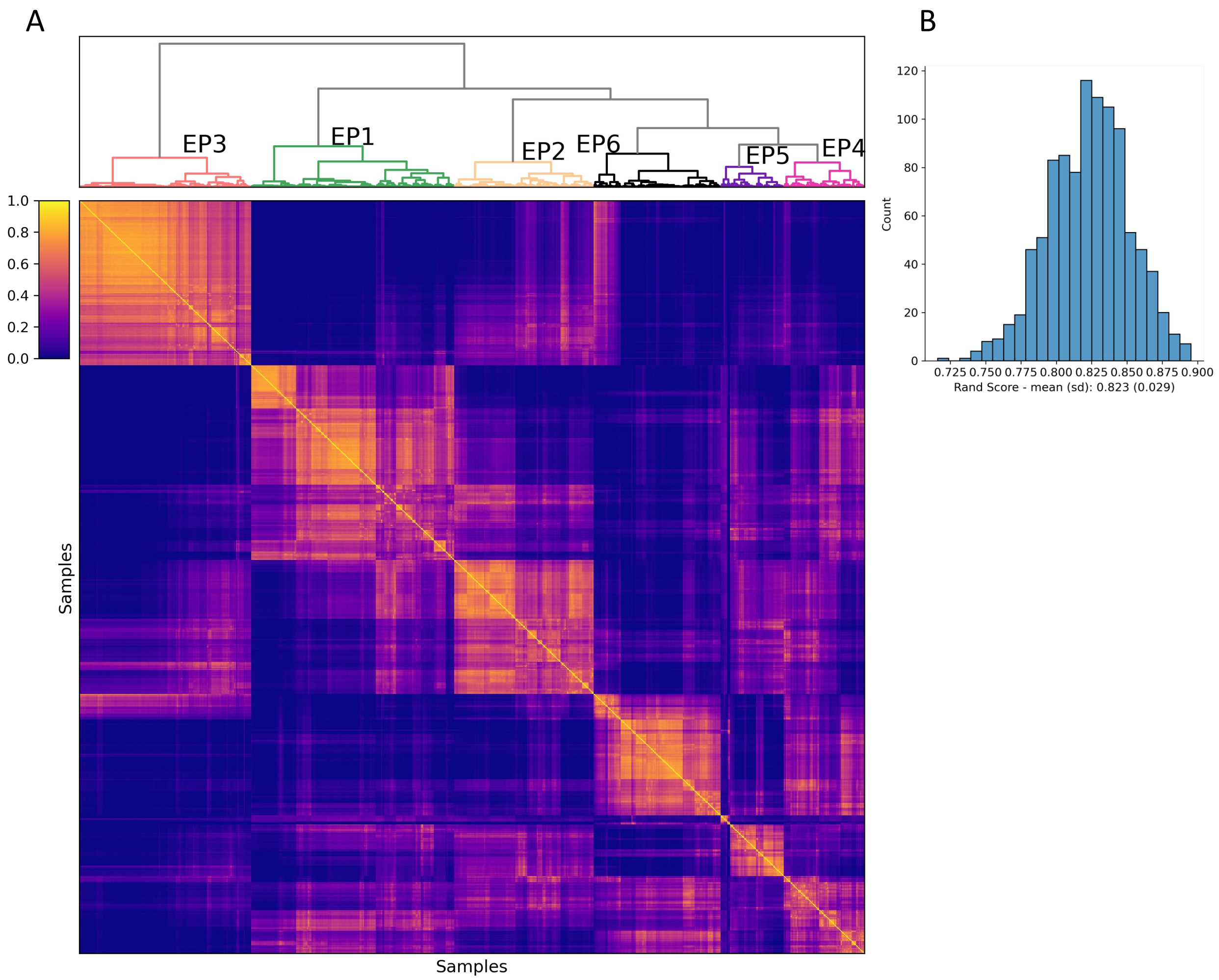


**Figure S2:** Endophenotypes of SARS-CoV-2 positive hospitalized patients (n=732) formed using consensus clustering of circulating proteome. A) A value in row i and column j of the heatmap shows the frequency that the ith patient and the jth patient were grouped together in the 1000 clusterings performed using 80% of randomly selected patients. Patients that are frequently grouped together form a final cluster. B) Histogram of Rand Index comparing each of the 1000 clusterings against the final consensus clustering. Rand Index is a measure of consistency between clusterings between 0 and 1, where 1 shows the highest degree of consistency (identical clusterings). The histogram shows a high degree of consistency.

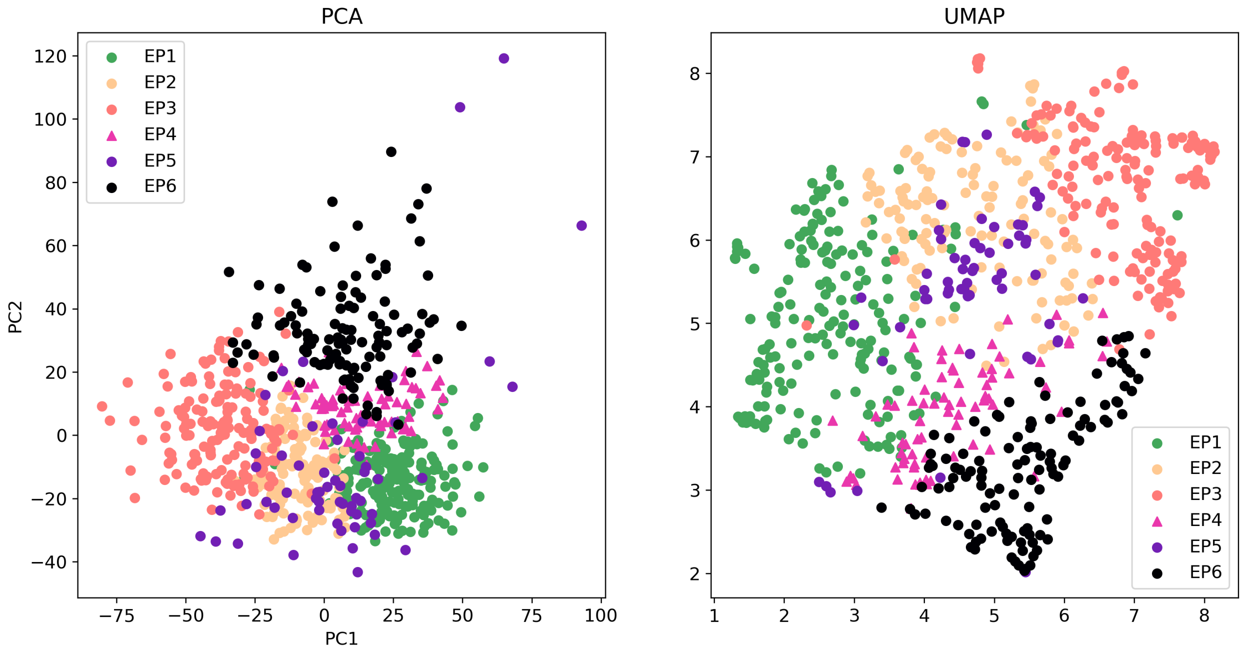


**Figure S3:** PCA and UMAP plots show the distribution of patients in each endophenotype in a two-dimensional figure. Sample colors are chosen to match those of Kaplan-Meier plots in Figure 2.

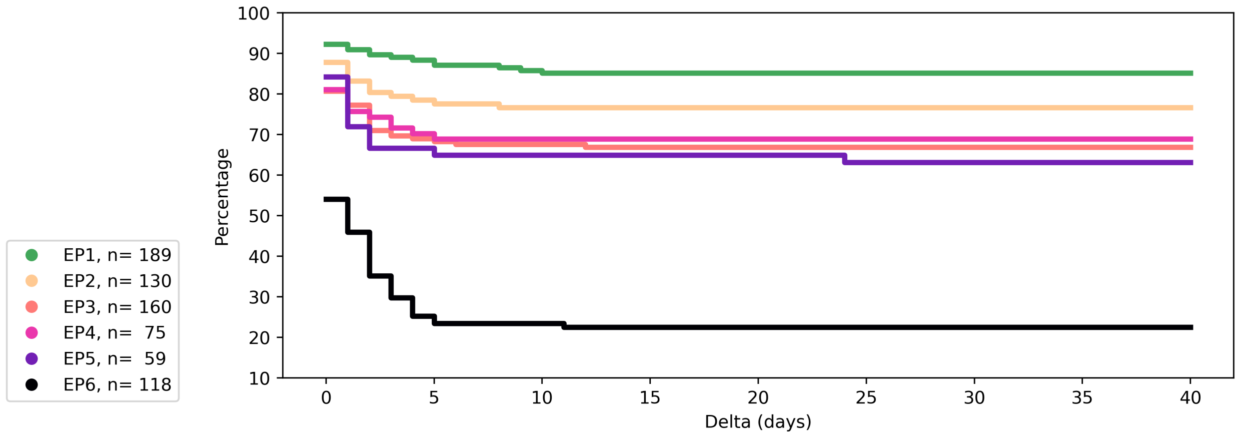


**Figure S4:** Kaplan-Meier analysis of the time between patients' admission to the hospital and their admission to ICU for each EP (Delta). Patients who died before admission to ICU are excluded from the analysis. A multivariate logrank test showed distinct patterns for each EP (P = 5.39E-30).

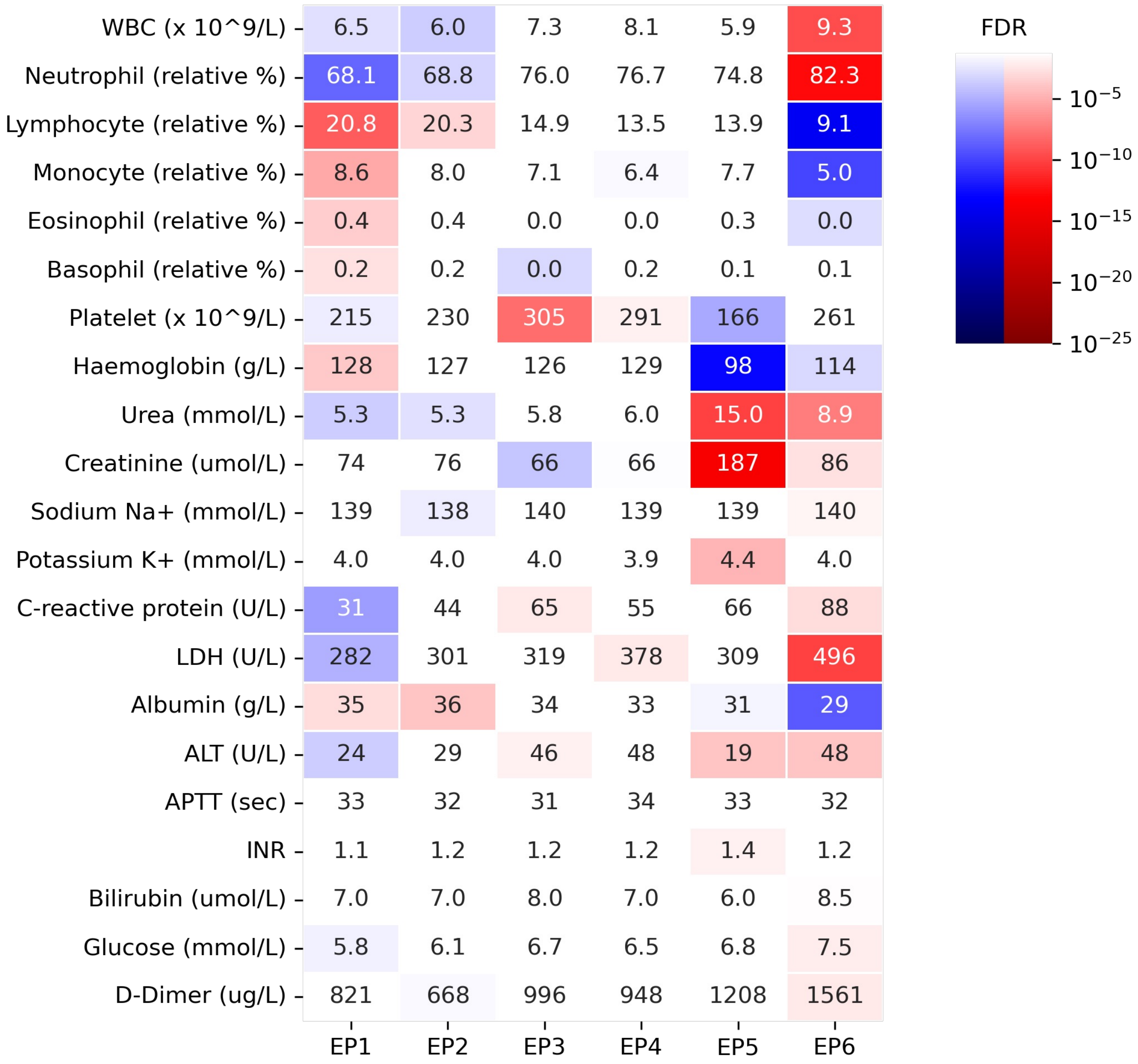


**Figure S5:** Patterns of blood markers for the first blood draw in the endophenotypes (EPs). Heatmaps show the false discovery rate (FDR) values for two-sided one-vs-rest Mann–Whitney U tests for 21 blood markers (first blood draw). FDR values below 0.05 are shown as white. The numerical values show the median value of the maker in each EP. Abbreviations used: WBC = white blood cells, LDH = lactate deshydrogenase, ALT = alanine aminotransferase, aPTT = activated partial thromboplastin time, INR = International Normalized Ratio.

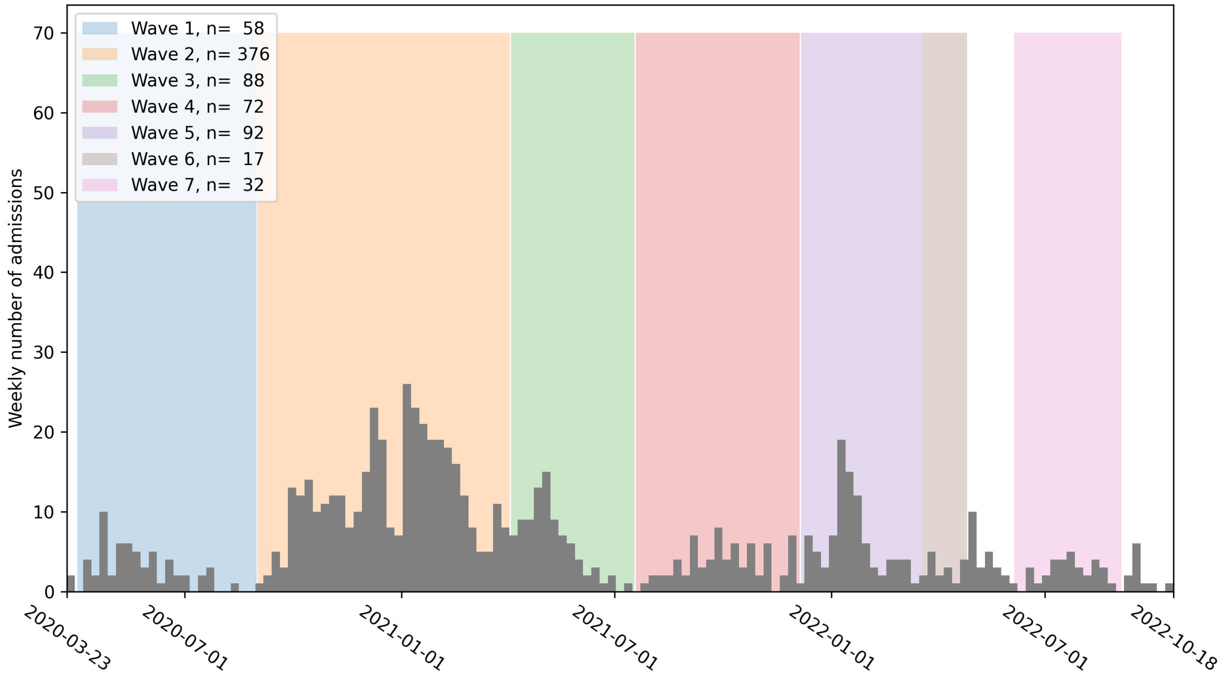


**Figure S6:** Weekly histogram of hospital admissions for patients used in the predictive model to define the predicted endophenotypes (PEPs) with known admission dates. Colors show different waves of COVID19 pandemic in Quebec according to National Institute of Public Health of Quebec (https://www.inspq.qc.ca/covid-19).

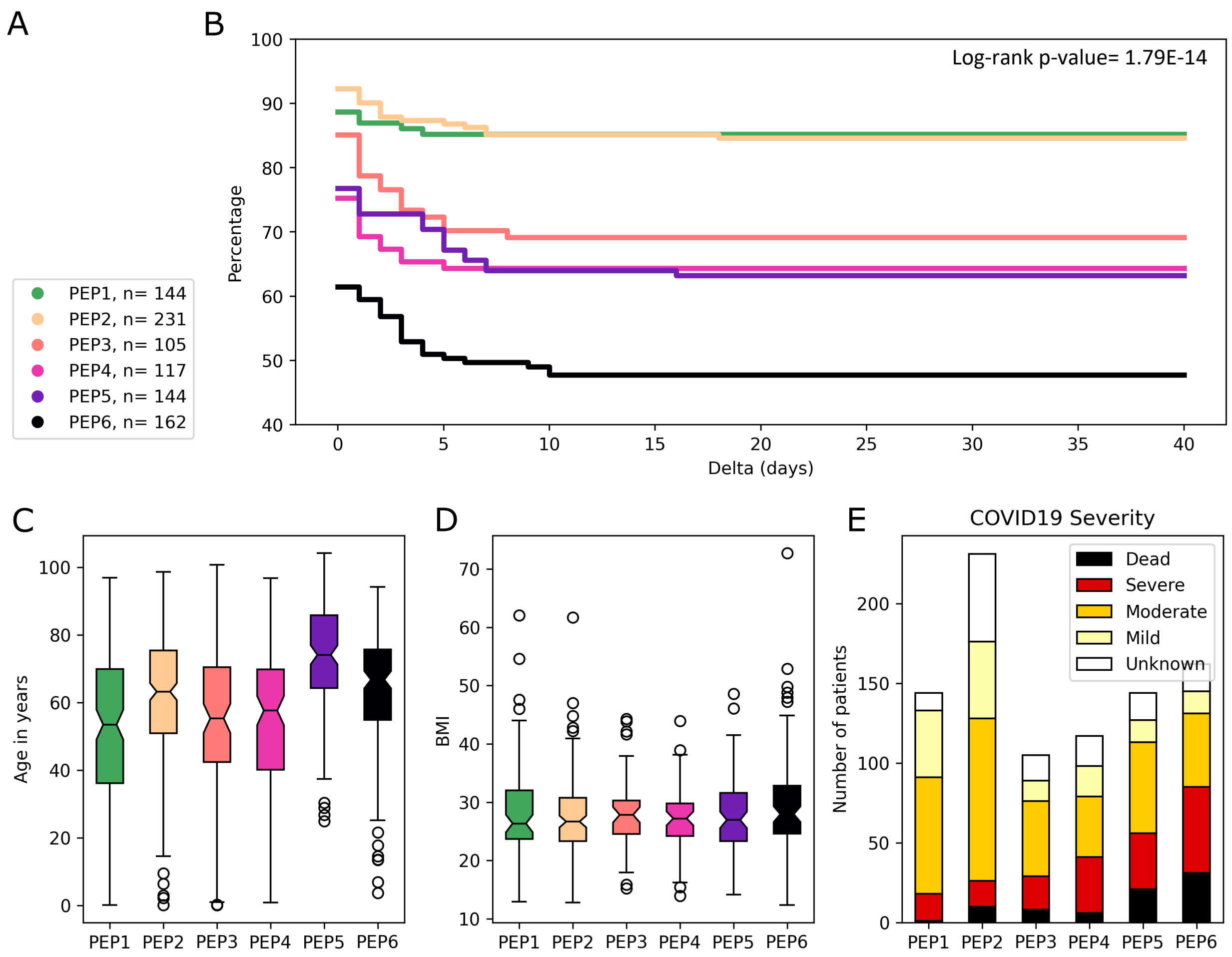


**Figure S7:** Characterization of predicted endophenotypes (PEPs) based on the prognostic model using 21 blood markers (first blood draw). A) The number of patients in each PEP and the colors used to represent them in panels B, C, D, and E. C) Kaplan-Meier analysis of the time between patients' admission to the hospital and their admission to intensive care unit (ICU) (or death if earlier) for each PEP (Delta). D) Distribution of age in each PEP. E) Distribution of BMI in each PEP. F) World health organization COVID-19 severity in each PEP.

**Supplementary Tables**

**Table S1:** Clinical and pathological characteristics of the patients and endophenotypes. The table is provided as a separate excel file.

**Table S2:** Enrichment of endophenotypes in different complications. The table is provided as a separate excel file.

**Table S3:** Characteristics of the endophenotypes based on blood markers and Roche diagnostic markers. The table is provided as a separate excel file.

**Table S4:** Characteristics of predicted endophenotypes (PEPs). The table is provided as a separate excel file.
